## Supplements 1 and 2 for "Vitamin D3 Supplementation at 5000 IU Daily for the Prevention of Influenza-Like Illness in Healthcare Workers: A Randomized Clinical Trial": S1 CONSORT Checklist.docx

| Section/Topic | Item No | Checklist item | Reported on page No |
| --- | --- | --- | --- |
| Title and abstract | | | |
|  | 1a | Identification as a randomized trial in the title | 1 |
|  | 1b | Structured summary of trial design, methods, results, and conclusions (for specific guidance see CONSORT for abstracts) | 2,3 |
| Introduction | | | |
| Background and objectives | 2a | Scientific background and explanation of rationale | 4-5 |
|  | 2b | Specific objectives or hypotheses | 5 |
| Methods | | | |
| Trial design | 3a | Description of trial design (such as parallel, factorial) including allocation ratio | 6-7, Figure 1 |
|  | 3b | Important changes to methods after trial commencement (such as eligibility criteria), with reasons | 6-7 |
| Participants | 4a | Eligibility criteria for participants | 6-7, Table 1 |
|  | 4b | Settings and locations where the data were collected | 1 |
| Interventions | 5 | The interventions for each group with sufficient details to allow replication, including how and when they were actually administered | 6-11 |
| Outcomes | 6a | Completely defined pre-specified primary and secondary outcome measures, including how and when they were assessed | 8-9 |
|  | 6b | Any changes to trial outcomes after the trial commenced, with reasons | 6-7 |
| Sample size | 7a | How sample size was determined | 7 |
|  | 7b | When applicable, explanation of any interim analyses and stopping guidelines | N/A |
| Randomization: |  |  |  |
| Sequence generation | 8a | Method used to generate the random allocation sequence | 6-7 |
|  | 8b | Type of randomization; details of any restriction (such as blocking and block size) | 6-7 |
| Allocation concealment mechanism | 9 | Mechanism used to implement the random allocation sequence (such as sequentially numbered containers), describing any steps taken to conceal the sequence until interventions were assigned | N/A |
| Implementation | 10 | Who generated the random allocation sequence, who enrolled participants, and who assigned participants to interventions | 6-7 |
| Blinding | 11a | If done, who was blinded after assignment to interventions (for example, participants, care providers, those assessing outcomes) and how | N/A |
|  | 11b | If relevant, description of the similarity of interventions | N/A |
| Statistical methods | 12a | Statistical methods used to compare groups for primary and secondary outcomes | 10-11 |
|  | 12b | Methods for additional analyses, such as subgroup analyses and adjusted analyses | 10-11 |
| Results | | | |
| Participant flow (a diagram is strongly recommended) | 13a | For each group, the numbers of participants who were randomly assigned, received intended treatment, and were analyzed for the primary outcome | 11-12, Figure 1 |
|  | 13b | For each group, losses and exclusions after randomization, together with reasons | Figure 1 |
| Recruitment | 14a | Dates defining the periods of recruitment and follow-up | 11-12 |
|  | 14b | Why the trial ended or was stopped | N/A |
| Baseline data | 15 | A table showing baseline demographic and clinical characteristics for each group | Table 2 |
| Numbers analysed | 16 | For each group, number of participants (denominator) included in each analysis and whether the analysis was by original assigned groups | 10-14, Figure 1 |
| Outcomes and estimation | 17a | For each primary and secondary outcome, results for each group, and the estimated effect size and its precision (such as 95% confidence interval) | 11-14, Figure 2, Table 3 |
|  | 17b | For binary outcomes, presentation of both absolute and relative effect sizes is recommended | Tables 3-4 |
| Ancillary analyses | 18 | Results of any other analyses performed, including subgroup analyses and adjusted analyses, distinguishing pre-specified from exploratory | 12-14, Table 4 |
| Harms | 19 | All important harms or unintended effects in each group (for specific guidance see CONSORT for harms) | 14-15, S2 |
| Discussion | | | |
| Limitations | 20 | Trial limitations, addressing sources of potential bias, imprecision, and, if relevant, multiplicity of analyses | 18-19 |
| Generalizability | 21 | Generalizability (external validity, applicability) of the trial findings | 19 |
| Interpretation | 22 | Interpretation consistent with results, balancing benefits and harms, and considering other relevant evidence | 16-19 |
| Other information | | |  |
| Registration | 23 | Registration number and name of trial registry | 6 |
| Protocol | 24 | Where the full trial protocol can be accessed, if available | Figshare |
| Funding | 25 | Sources of funding and other support (such as supply of drugs), role of funders | 1, 7 |
