## Supplements 1 and 2 for "Vitamin D3 Supplementation at 5000 IU Daily for the Prevention of Influenza-Like Illness in Healthcare Workers: A Randomized Clinical Trial": S2 Adverse Events.docx

**S2. Adverse events (AE) and relatedness among intervention group during 9-month study period**

| **Adverse events (AE) and relatedness among intervention group during 9-month study period** | | | | |
| --- | --- | --- | --- | --- |
| **Adverse Events = 388** | **AE (%)** | Probably Related | Possibly Related | Unrelated |
| Fatigue | **36 (9.3%)** | 0 | 4 | 32 |
| Urinary frequency | **24 (6.2%)** | 1 | 7 | 16 |
| Nausea | **23 (5.9%)** | 0 | 8 | 15 |
| Lower back pain | **21 (5.4%)** | 0 | 2 | 19 |
| Thirst/dry mouth | **17 (4.4%)** | 1 | 5 | 11 |
| Abdominal pain | **11 (2.8%)** | 1 | 4 | 6 |
| Back pain | **11 (2.8%)** | 0 | 1 | 10 |
| Palpitations | **10 (2.6%)** | 0 | 5 | 5 |
| Stomach upset | **10 (2.6%)** | 0 | 4 | 6 |
| Urinary tract infection | **9 (2.3%)** | 0 | 1 | 8 |
| Headache | **8 (2.1%)** | 0 | 0 | 8 |
| GERD | **7 (1.8%)** | 0 | 1 | 6 |
| Depression | **6 (1.5%)** | 0 | 1 | 5 |
| Lack of energy | **6 (1.5%)** | 0 | 1 | 5 |
| Anxiety | **5 (1.3%)** | 0 | 0 | 5 |
| Bone pain | **5 (1.3%)** | 0 | 0 | 5 |
| Constipation | **5 (1.3%)** | 0 | 4 | 1 |
| High blood pressure | **5 (1.3%)** | 0 | 1 | 4 |
| Muscle weakness | **5 (1.3%)** | 0 | 0 | 5 |
| Anxiety/depression | **4 (1.0%)** | 0 | 0 | 4 |
| Sinus infection/sinusitis | **4 (1.0%)** | 0 | 0 | 4 |
| Joint pain | **3 (0.8%)** | 0 | 0 | 3 |
| Rash | **3 (0.8%)** | 0 | 1 | 2 |
| Stomach upset/nausea | **3 (0.8%)** | 0 | 1 | 2 |
| Stomach virus | **3 (0.8%)** | 0 | 0 | 3 |
| Strong heartbeat | **3 (0.8%)** | 0 | 2 | 1 |
| Vomiting | **3 (0.8%)** | 0 | 1 | 2 |
| Bronchitis | **2 (0.5%)** | 0 | 0 | 2 |
| Dog bite | **2 (0.5%)** | 0 | 0 | 2 |
| Hives | **2 (0.5%)** | 0 | 1 | 1 |
| Lightheadedness | **2 (0.5%)** | 0 | 0 | 2 |
| Sciatica | **2 (0.5%)** | 0 | 0 | 2 |
| Tooth infection | **2 (0.5%)** | 0 | 0 | 2 |
| Upper back pain | **2 (0.5%)** | 0 | 1 | 1 |
| Weight gain | **2 (0.5%)** | 0 | 0 | 2 |
| Abdominal cramping/loose stools | **1 (0.3%)** | 0 | 1 | 0 |
| Abdominal muscle pain | **1 (0.3%)** | 0 | 0 | 1 |
| Abdominal/bloating | **1 (0.3%)** | 0 | 0 | 1 |
| Anemia | **1 (0.3%)** | 0 | 0 | 1 |
| Angioedema | **1 (0.3%)** | 0 | 0 | 1 |
| Attention deficit | **1 (0.3%)** | 0 | 0 | 1 |
| Bad taste in mouth | **1 (0.3%)** | 0 | 0 | 1 |
| Bloating, sense of fullness | **1 (0.3%)** | 0 | 0 | 1 |
| Body aches, joint aches, pain in legs and knees | **1 (0.3%)** | 0 | 0 | 1 |
| Bone and joint pain | **1 (0.3%)** | 0 | 1 | 0 |
| Bone pain-mild joint aches and mild post exercise joint discomfort. | **1 (0.3%)** | 0 | 0 | 1 |
| Bright red blood per rectum | **1 (0.3%)** | 0 | 0 | 1 |
| Burning sensation arms and legs | **1 (0.3%)** | 1 | 0 | 0 |
| Cellulitis | **1 (0.3%)** | 0 | 0 | 1 |
| Chest Pain and palpitations | **1 (0.3%)** | 0 | 1 | 0 |
| Confusion | **1 (0.3%)** | 0 | 0 | 1 |
| Conjunctivitis of the left eye | **1 (0.3%)** | 0 | 0 | 1 |
| Consistent severe itching of right inner ear | **1 (0.3%)** | 0 | 0 | 1 |
| Cramping and soreness in both legs | **1 (0.3%)** | 0 | 1 | 0 |
| Degenerative disc disease | **1 (0.3%)** | 0 | 0 | 1 |
| Diabetes type II | **1 (0.3%)** | 0 | 0 | 1 |
| Diarrhea | **1 (0.3%)** | 0 | 0 | 1 |
| Dizziness | **1 (0.3%)** | 0 | 0 | 1 |
| Fascia tightness/stiffness-neck, shoulders, back, arms, legs, feet | **1 (0.3%)** | 0 | 0 | 1 |
| Feeling hot | **1 (0.3%)** | 0 | 0 | 1 |
| Fibromyalgia | **1 (0.3%)** | 0 | 0 | 1 |
| Flank pain | **1 (0.3%)** | 0 | 0 | 1 |
| Fluid retention | **1 (0.3%)** | 0 | 0 | 1 |
| Food poisoning | **1 (0.3%)** | 0 | 0 | 1 |
| Foot injury from running | **1 (0.3%)** | 0 | 0 | 1 |
| Gallbladder polyps | **1 (0.3%)** | 0 | 0 | 1 |
| Ganglion cyst 3rd MCP and index finger right hand | **1 (0.3%)** | 0 | 0 | 1 |
| Gastroparesis | **1 (0.3%)** | 0 | 0 | 1 |
| GI upset | **1 (0.3%)** | 0 | 1 | 0 |
| GI virus | **1 (0.3%)** | 0 | 0 | 1 |
| Gum infection | **1 (0.3%)** | 0 | 0 | 1 |
| H. pylori | **1 (0.3%)** | 0 | 0 | 1 |
| Hair loss | **1 (0.3%)** | 0 | 0 | 1 |
| Hand pain | **1 (0.3%)** | 0 | 0 | 1 |
| Having a bowel movement more often | **1 (0.3%)** | 0 | 1 | 0 |
| Heaviness in lower extremities | **1 (0.3%)** | 0 | 0 | 1 |
| Hematemesis | **1 (0.3%)** | 0 | 0 | 1 |
| High heartbeat | **1 (0.3%)** | 0 | 0 | 1 |
| Hypersensitivity to sun | **1 (0.3%)** | 1 | 0 | 0 |
| Increase in occurrence of pain in right hip and right knee | **1 (0.3%)** | 0 | 0 | 1 |
| Increased drinking and urination | **1 (0.3%)** | 0 | 1 | 0 |
| Increased frequency of IBS pain | **1 (0.3%)** | 0 | 0 | 1 |
| Infected ingrown toenail | **1 (0.3%)** | 0 | 0 | 1 |
| Infected R toe | **1 (0.3%)** | 0 | 0 | 1 |
| Insomnia-intermittent | **1 (0.3%)** | 0 | 0 | 1 |
| Jaw pain | **1 (0.3%)** | 0 | 1 | 0 |
| Kidney stone | **1 (0.3%)** | 0 | 0 | 1 |
| Knee pain | **1 (0.3%)** | 0 | 0 | 1 |
| Knee pain and shin pain | **1 (0.3%)** | 0 | 1 | 0 |
| Labral tear in hip joint | **1 (0.3%)** | 0 | 0 | 1 |
| Left knee pain | **1 (0.3%)** | 0 | 0 | 1 |
| Low B12 | **1 (0.3%)** | 0 | 0 | 1 |
| Low back pain-intermittent | **1 (0.3%)** | 0 | 0 | 1 |
| Low iron | **1 (0.3%)** | 0 | 0 | 1 |
| Memory issue | **1 (0.3%)** | 0 | 0 | 1 |
| Menstrual issue | **1 (0.3%)** | 0 | 0 | 1 |
| More frequent urination and urge | **1 (0.3%)** | 0 | 0 | 1 |
| Mouth lesions | **1 (0.3%)** | 1 | 0 | 0 |
| Muscle pain | **1 (0.3%)** | 0 | 0 | 1 |
| Muscle pain in arms | **1 (0.3%)** | 0 | 0 | 1 |
| Muscle spasms for 2-3 days, both sides of rib cage | **1 (0.3%)** | 0 | 0 | 1 |
| Neck pain | **1 (0.3%)** | 0 | 0 | 1 |
| Nodules on right lower forearm and right foot near ankle | **1 (0.3%)** | 0 | 0 | 1 |
| Nodules on throat | **1 (0.3%)** | 0 | 0 | 1 |
| Occasional stomach pains | **1 (0.3%)** | 0 | 0 | 1 |
| Oral tenderness | **1 (0.3%)** | 1 | 0 | 0 |
| Otitis externa | **1 (0.3%)** | 0 | 0 | 1 |
| Pain in fingers-possibly arthritis | **1 (0.3%)** | 0 | 0 | 1 |
| Pain-knee | **1 (0.3%)** | 0 | 0 | 1 |
| Pelvic pain | **1 (0.3%)** | 0 | 0 | 1 |
| Pulled muscle left elbow | **1 (0.3%)** | 0 | 0 | 1 |
| PVCs | **1 (0.3%)** | 0 | 0 | 1 |
| Rapid heartbeat | **1 (0.3%)** | 0 | 1 | 0 |
| Raynaud's | **1 (0.3%)** | 0 | 0 | 1 |
| Recurrent strep throat infections | **1 (0.3%)** | 0 | 0 | 1 |
| Rib pain | **1 (0.3%)** | 0 | 1 | 0 |
| Ridges in nails | **1 (0.3%)** | 0 | 0 | 1 |
| Right ankle, left knee and left wrist worsened pain | **1 (0.3%)** | 0 | 0 | 1 |
| Right big toe swollen/stiff | **1 (0.3%)** | 0 | 0 | 1 |
| Right knee pain | **1 (0.3%)** | 0 | 0 | 1 |
| Right leg pain | **1 (0.3%)** | 0 | 0 | 1 |
| Right ovarian cyst | **1 (0.3%)** | 0 | 0 | 1 |
| Right shoulder pain | **1 (0.3%)** | 0 | 0 | 1 |
| Right shoulder stiffening | **1 (0.3%)** | 0 | 0 | 1 |
| Ruptured Meckel's diverticulum | **1 (0.3%)** | 0 | 0 | 1 |
| Scabies | **1 (0.3%)** | 0 | 0 | 1 |
| Shingles | **1 (0.3%)** | 0 | 0 | 1 |
| Short moments of forgetting things | **1 (0.3%)** | 0 | 0 | 1 |
| Shoulder pain-frozen shoulder | **1 (0.3%)** | 0 | 0 | 1 |
| Shoulder surgery | **1 (0.3%)** | 0 | 0 | 1 |
| Spasms in shoulder resulting from surgery | **1 (0.3%)** | 0 | 0 | 1 |
| Stomach cramping | **1 (0.3%)** | 0 | 0 | 1 |
| Stomach pain | **1 (0.3%)** | 0 | 0 | 1 |
| Stomach pain intermittent | **1 (0.3%)** | 0 | 1 | 0 |
| Stomach upset and constipation - intermittent | **1 (0.3%)** | 0 | 0 | 1 |
| Stye in left eye | **1 (0.3%)** | 0 | 0 | 1 |
| Swelling of feet, weakness in hands, discomfort in hands and feet | **1 (0.3%)** | 0 | 0 | 1 |
| Swollen gland near jaw | **1 (0.3%)** | 0 | 0 | 1 |
| Swollen gum | **1 (0.3%)** | 0 | 0 | 1 |
| Tachycardia and chest pain thinks it was indigestion | **1 (0.3%)** | 0 | 1 | 0 |
| Tachycardia and pressure in chest | **1 (0.3%)** | 0 | 0 | 1 |
| Thyroid antibodies elevated | **1 (0.3%)** | 0 | 0 | 1 |
| Thyroidectomy | **1 (0.3%)** | 0 | 0 | 1 |
| Tick bite | **1 (0.3%)** | 0 | 0 | 1 |
| TMJ | **1 (0.3%)** | 0 | 1 | 0 |
| Toes, and fingers twitching, muscle twitching | **1 (0.3%)** | 0 | 0 | 1 |
| Unable to catch breath | **1 (0.3%)** | 0 | 0 | 1 |
| Unknown allergic reaction | **1 (0.3%)** | 0 | 0 | 1 |
| Upper back pain and Upper leg pain | **1 (0.3%)** | 0 | 0 | 1 |
| Urinary urgency | **1 (0.3%)** | 0 | 0 | 1 |
| Vaginal burning | **1 (0.3%)** | 0 | 0 | 1 |
| Vertigo | **1 (0.3%)** | 0 | 0 | 1 |
| Worsened acne | **1 (0.3%)** | 0 | 1 | 0 |
| Worsened neck, shoulder, back stiffness and soreness | **1 (0.3%)** | 0 | 0 | 1 |
| Worsening back and right leg pain | **1 (0.3%)** | 1 | 0 | 0 |
| Wound infection | **1 (0.3%)** | 0 | 0 | 1 |
| Yeast infections | **1 (0.3%)** | 0 | 0 | 1 |
| **Total Adverse Events (AEs)** | **100%** | **8** | **71** | **309** |
